# Cost-Effectiveness of Expanding Housing Access for People Experiencing Homelessness and Opioid Use Disorder in California

**DOI:** 10.64898/2026.10.15.26365505

**Authors:** Yuewei Ling, Sean Tsung, Fernando Alarid-Escudero, Jeremy D. Goldhaber-Fiebert, Margaret L. Brandeau, Marissa B. Reitsma

**Author notes:** Corresponding author. Address: Department of Management Science and Engineering, Huang Engineering Center, 475 via Ortega, Stanford, CA 94305., Yuewei Ling. Contributed equally (Yuewei Ling, Sean Tsung). **Contact Information:**; Sean Tsung; Fernando Alarid-Escudero; Jeremy D. Goldhaber-Fiebert; Margaret L. Brandeau, Marissa B. Reitsma.

## Abstract

People experiencing homelessness (PEH) have substantially lower quality-adjusted life expectancy and substantially higher healthcare spending compared to housed populations. California has the largest population of PEH in the United States, and an estimated ten percent of PEH have co-occurring opioid use disorder (OUD). We developed a lifetime, individual-level microsimulation of the 18,928 PEH with OUD in California to evaluate the health and economic effects of expanding access to housing through a Housing First approach, which does not require OUD treatment or abstinence. Increases in permanent housing and shelter access were based on Los Angeles County’s Measure A goals, and we accounted for both housing and healthcare costs. Expanding housing access increased mean age at death by 0.34 years, reduced fatal overdoses for the cohort by 39, and produced an incremental cost-effectiveness ratio of $31,083 per quality-adjusted life-year gained, well below conventional willingness-to-pay thresholds. We provide evidence that housing is a high-value health intervention for PEH with OUD in California. Policymakers should support Housing First programs as part of integrated responses to the intertwined crises of homelessness and opioid use.

## INTRODUCTION

Homelessness remains at historically high levels in the United States, with nearly 750,000 people estimated to be experiencing homelessness on a given night during 2025 (about 0.2% of the population).^1^ Mortality rates among people experiencing homelessness (PEH) are high,^2, 3^ with opioid overdose being the leading cause of death among PEH.^2–6^ These challenges are especially urgent in California, which has an estimated 182,000 PEH, the largest of any state.^1^ In a California survey, 78% of adult PEH reported spending most nights in the past six months unsheltered, living in places not meant for human habitation; 10.4% of PEH reported regular opioid use during that period, often in combination with methamphetamines, which increases overdose risk.^7, 8^

The risks associated with opioid use disorder (OUD) are more severe among PEH than among housed individuals due to unsafe living conditions, limited access to healthcare, and co-occurring mental health conditions. As a result, PEH have disproportionately higher rates of overdose, unmet behavioral health needs, inadequate access to treatment, and barriers to sustained recovery.^7^ Barriers to OUD treatment include lack of stable housing, stigma, fragmented care, and practical challenges such as transportation and medication security.^9–11^ Furthermore, compared to sheltered PEH, unsheltered PEH have higher rates of substance use, overdose, morbidity, and mortality, and face greater barriers to care.^7, 12^

Despite increasing attention to both homelessness and addiction, policy responses often remain fragmented, with housing and healthcare financed, delivered, and evaluated through separate systems.^13, 14^At present, evidence remains limited to guide local, integrated policies that address both housing instability and addiction care. Previous economic evaluations have examined housing interventions and OUD treatments separately,^15, 16^ but relatively few have jointly evaluated housing provision and OUD treatment as coordinated intervention components among PEH.^17, 18^ Furthermore, stratification by shelter status is also essential for accurate risk assessment, service planning, and intervention design.^12, 19^ A prior analysis used a dynamic model to assess health and economic outcomes of housing provision for PEH with OUD, finding it to be cost-effective.^20^ That analysis considered a representative national cohort and characterized housing status as either housed or unhoused, without distinguishing between sheltered and unsheltered PEH. Under the housing intervention, all individuals were provided stable housing at the start of the modeled time horizon, with subsequent housing loss allowed but without modeling subsequent transitions back into housing.

Drawing upon recent evidence from the California Statewide Study of People Experiencing Homelessness,^8^ we developed a microsimulation model to evaluate whether expanding access to temporary shelter and permanent housing for PEH with OUD can improve health outcomes and represent good value for public spending in California.

## METHODS

### Overview

We developed an individual-level, discrete-time microsimulation model that tracks monthly transitions of people with OUD across three housing states (unsheltered, sheltered, and permanently housed) and three opioid use states (out of treatment, in treatment, and abstinent). The monthly cycle length reflects the rapid transitions observed in housing instability and opioid use status. We modeled a cohort of 18,928 PEH with OUD, which represented the estimated population of PEH with OUD in California. Model inputs (**Table 1**) were informed by a combination of California data and published literature on mortality, overdose risk, OUD treatment, costs, and health-related quality of life. Initial OUD population characteristics (distributions of age, sex, and housing states) were estimated based on recent studies in California.^7, 8^

**Table 1.**
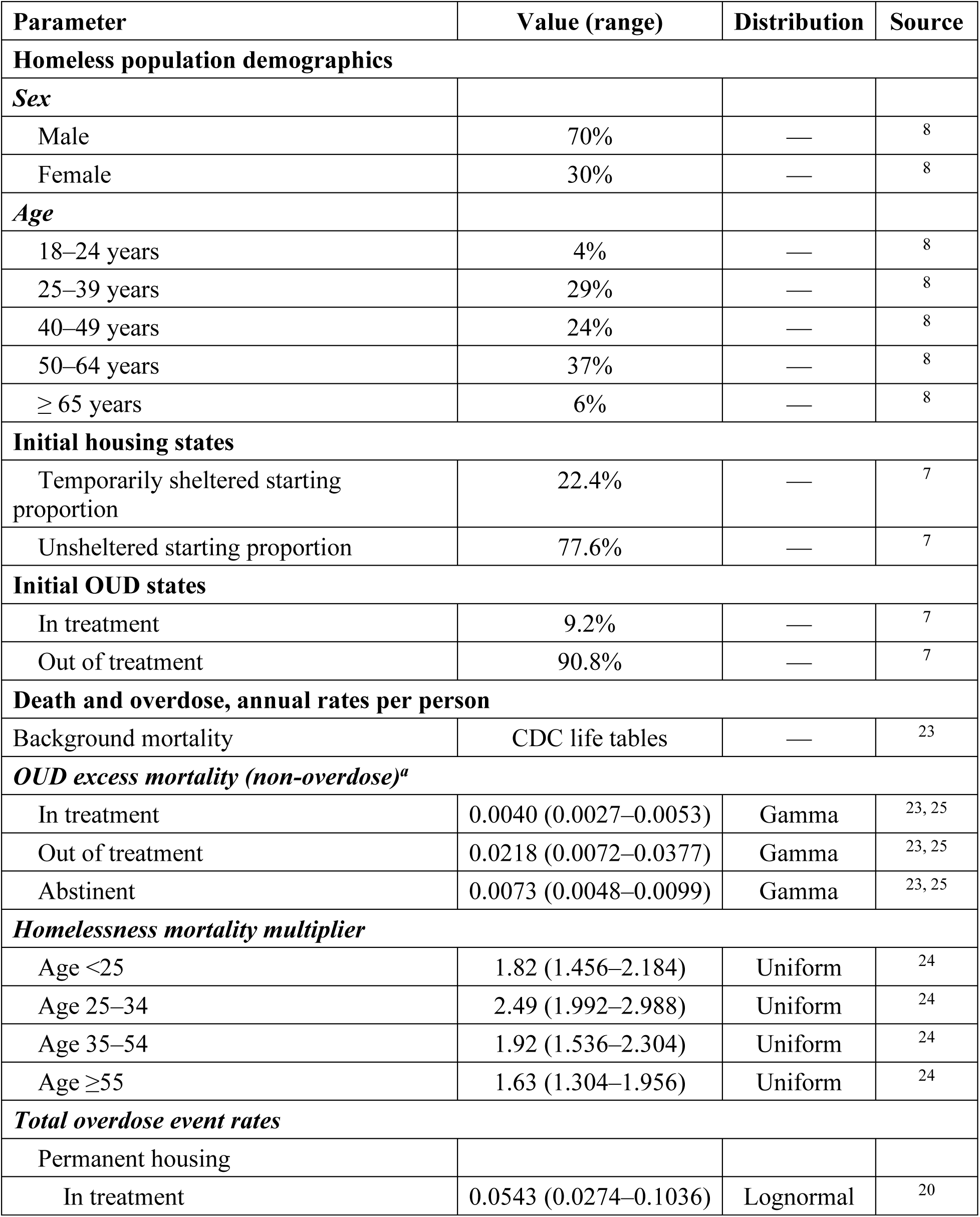
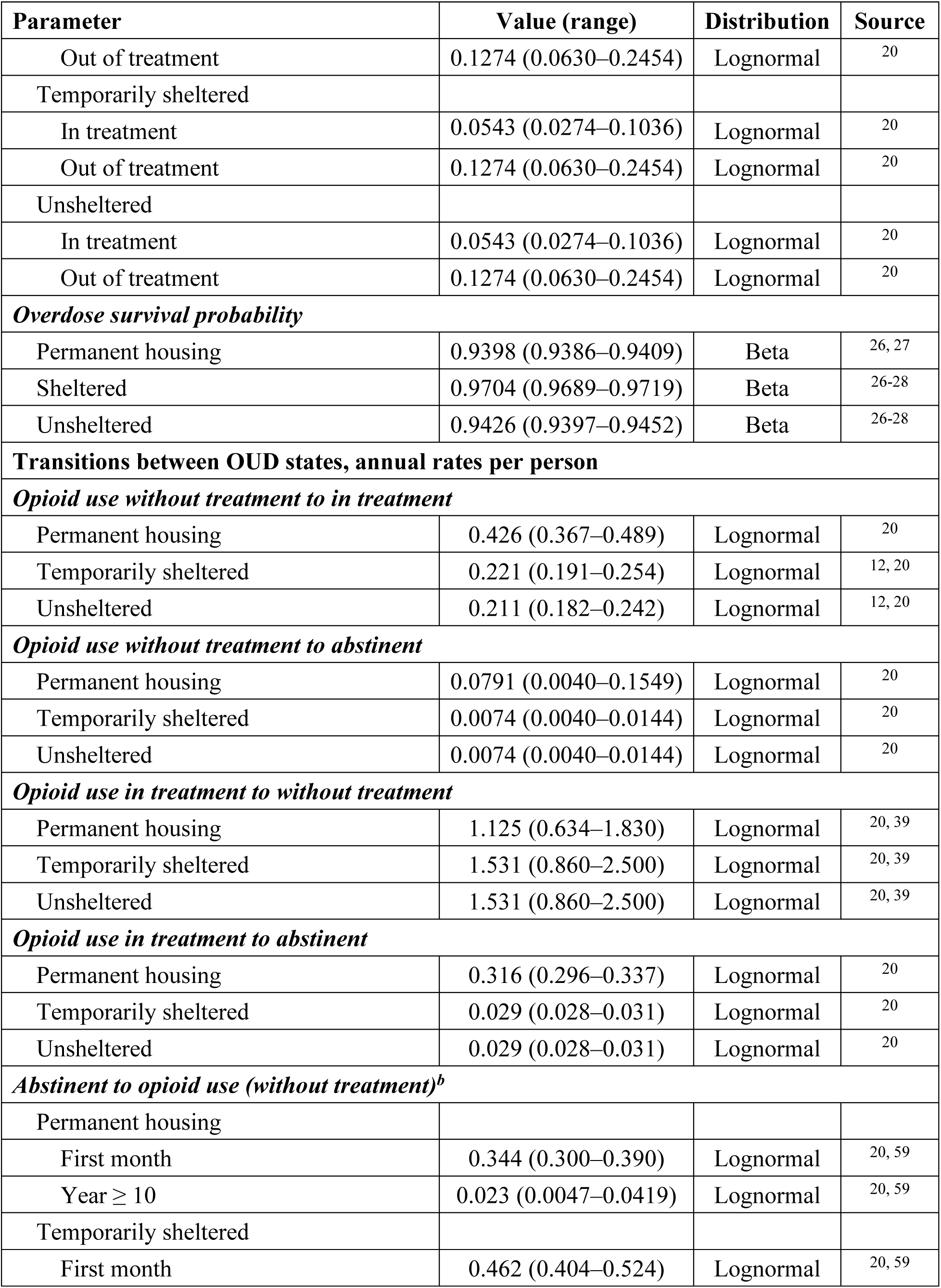
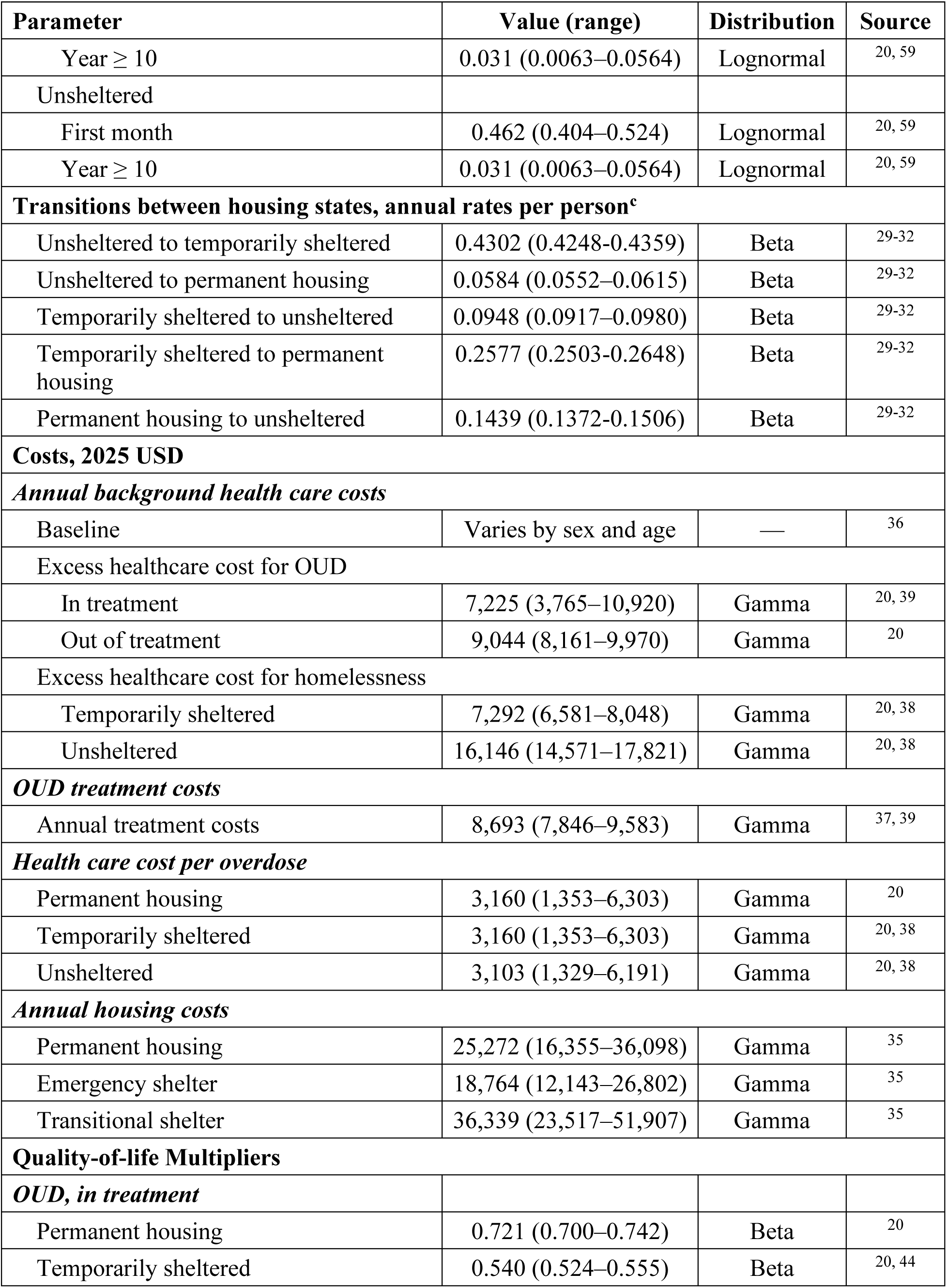
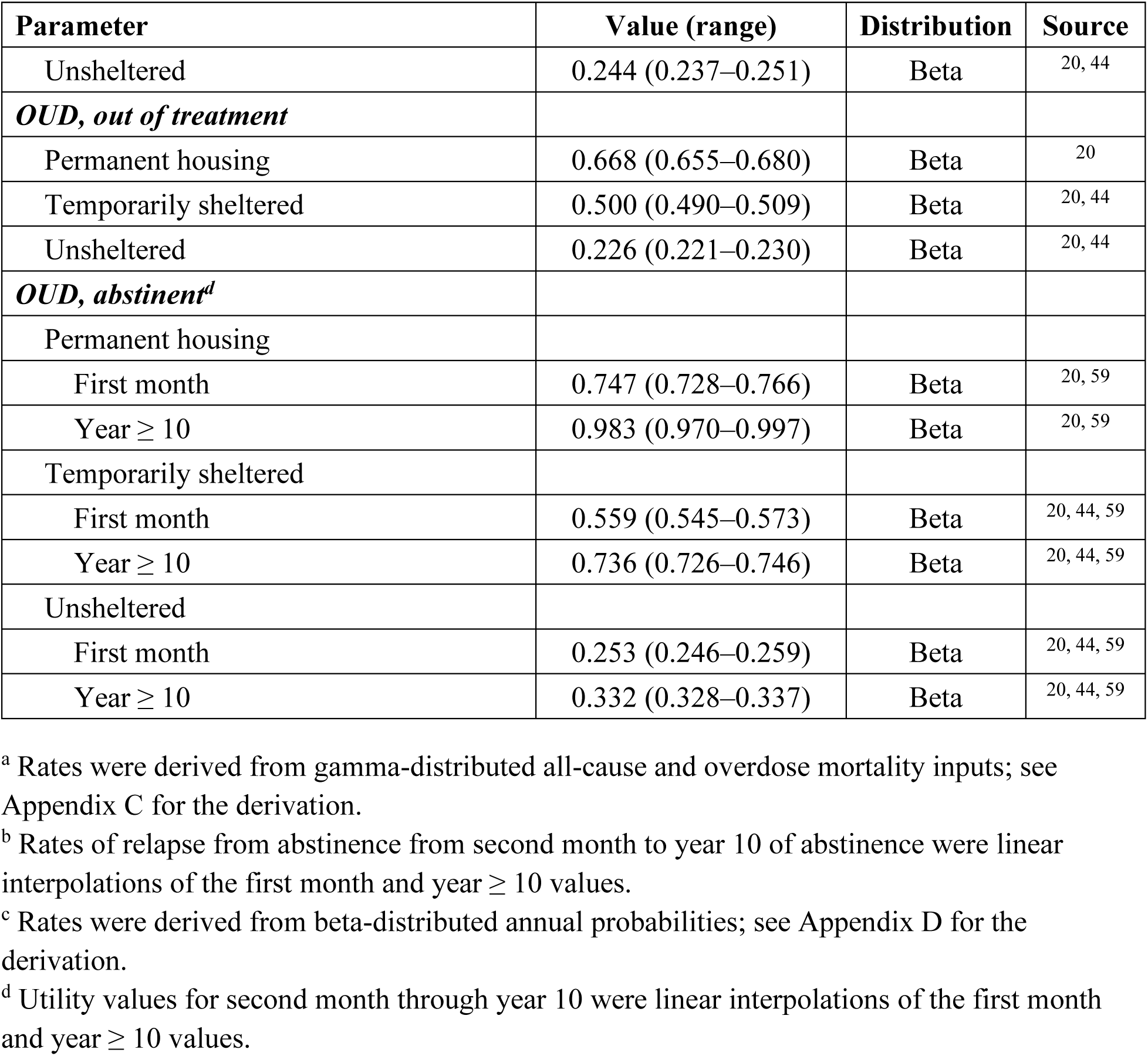
Base Case Parameter Values and Sources.

| Parameter | Value (range) | Distribution | Source |
| --- | --- | --- | --- |
| **Homeless population demographics** |  |  |  |
| *Sex* |  |  |  |
| Male | 70% | — | 8 |
| Female | 30% | — | 8 |
| *Age* |  |  |  |
| 18–24 years | 4% | — | 8 |
| 25–39 years | 29% | — | 8 |
| 40–49 years | 24% | — | 8 |
| 50–64 years | 37% | — | 8 |
| ≥ 65 years | 6% | — | 8 |
| **Initial housing states** |  |  |  |
| Temporarily sheltered starting proportion | 22.4% | — | 7 |
| Unsheltered starting proportion | 77.6% | — | 7 |
| **Initial OUD states** |  |  |  |
| In treatment | 9.2% | — | 7 |
| Out of treatment | 90.8% | — | 7 |
| **Death and overdose, annual rates per person** |  |  |  |
| Background mortality | CDC life tables | — | 23 |
| ***OUD excess mortality (non-overdose)<sup>a</sup>*** |  |  |  |
| In treatment | 0.0040 (0.0027–0.0053) | Gamma | 23, 25 |
| Out of treatment | 0.0218 (0.0072–0.0377) | Gamma | 23, 25 |
| Abstinent | 0.0073 (0.0048–0.0099) | Gamma | 23, 25 |
| ***Homelessness mortality multiplier*** |  |  |  |
| Age <25 | 1.82 (1.456–2.184) | Uniform | 24 |
| Age 25–34 | 2.49 (1.992–2.988) | Uniform | 24 |
| Age 35–54 | 1.92 (1.536–2.304) | Uniform | 24 |
| Age ≥55 | 1.63 (1.304–1.956) | Uniform | 24 |
| ***Total overdose event rates*** |  |  |  |
| Permanent housing |  |  |  |
| In treatment | 0.0543 (0.0274–0.1036) | Lognormal | 20 |
| Out of treatment | 0.1274 (0.0630–0.2454) | Lognormal | 20 |
| Temporarily sheltered |  |  |  |
| In treatment | 0.0543 (0.0274–0.1036) | Lognormal | 20 |
| Out of treatment | 0.1274 (0.0630–0.2454) | Lognormal | 20 |
| Unsheltered |  |  |  |
| In treatment | 0.0543 (0.0274–0.1036) | Lognormal | 20 |
| Out of treatment | 0.1274 (0.0630–0.2454) | Lognormal | 20 |
| ***Overdose survival probability*** |  |  |  |
| Permanent housing | 0.9398 (0.9386–0.9409) | Beta | 26, 27 |
| Sheltered | 0.9704 (0.9689–0.9719) | Beta | 26-28 |
| Unsheltered | 0.9426 (0.9397–0.9452) | Beta | 26-28 |
| **Transitions between OUD states, annual rates per person** |  |  |  |
| ***Opioid use without treatment to in treatment*** |  |  |  |
| Permanent housing | 0.426 (0.367–0.489) | Lognormal | 20 |
| Temporarily sheltered | 0.221 (0.191–0.254) | Lognormal | 12, 20 |
| Unsheltered | 0.211 (0.182–0.242) | Lognormal | 12, 20 |
| ***Opioid use without treatment to abstinent*** |  |  |  |
| Permanent housing | 0.0791 (0.0040–0.1549) | Lognormal | 20 |
| Temporarily sheltered | 0.0074 (0.0040–0.0144) | Lognormal | 20 |
| Unsheltered | 0.0074 (0.0040–0.0144) | Lognormal | 20 |
| ***Opioid use in treatment to without treatment*** |  |  |  |
| Permanent housing | 1.125 (0.634–1.830) | Lognormal | 20, 39 |
| Temporarily sheltered | 1.531 (0.860–2.500) | Lognormal | 20, 39 |
| Unsheltered | 1.531 (0.860–2.500) | Lognormal | 20, 39 |
| ***Opioid use in treatment to abstinent*** |  |  |  |
| Permanent housing | 0.316 (0.296–0.337) | Lognormal | 20 |
| Temporarily sheltered | 0.029 (0.028–0.031) | Lognormal | 20 |
| Unsheltered | 0.029 (0.028–0.031) | Lognormal | 20 |
| ***Abstinent to opioid use (without treatment)<sup>b</sup>*** |  |  |  |
| Permanent housing |  |  |  |
| First month | 0.344 (0.300–0.390) | Lognormal | 20, 59 |
| Year ≥ 10 | 0.023 (0.0047–0.0419) | Lognormal | 20, 59 |
| Temporarily sheltered |  |  |  |
| First month | 0.462 (0.404–0.524) | Lognormal | 20, 59 |
| Year $\geq 10$ | 0.031 (0.0063–0.0564) | Lognormal | 20, 59 |
| Unsheltered |  |  |  |
| First month | 0.462 (0.404–0.524) | Lognormal | 20, 59 |
| Year $\geq 10$ | 0.031 (0.0063–0.0564) | Lognormal | 20, 59 |
| **Transitions between housing states, annual rates per person<sup>c</sup>** |  |  |  |
| Unsheltered to temporarily sheltered | 0.4302 (0.4248–0.4359) | Beta | 29–32 |
| Unsheltered to permanent housing | 0.0584 (0.0552–0.0615) | Beta | 29–32 |
| Temporarily sheltered to unsheltered | 0.0948 (0.0917–0.0980) | Beta | 29–32 |
| Temporarily sheltered to permanent housing | 0.2577 (0.2503–0.2648) | Beta | 29–32 |
| Permanent housing to unsheltered | 0.1439 (0.1372–0.1506) | Beta | 29–32 |
| **Costs, 2025 USD** |  |  |  |
| *Annual background health care costs* |  |  |  |
| Baseline | Varies by sex and age | — | 36 |
| Excess healthcare cost for OUD |  |  |  |
| In treatment | 7,225 (3,765–10,920) | Gamma | 20, 39 |
| Out of treatment | 9,044 (8,161–9,970) | Gamma | 20 |
| Excess healthcare cost for homelessness |  |  |  |
| Temporarily sheltered | 7,292 (6,581–8,048) | Gamma | 20, 38 |
| Unsheltered | 16,146 (14,571–17,821) | Gamma | 20, 38 |
| *OUD treatment costs* |  |  |  |
| Annual treatment costs | 8,693 (7,846–9,583) | Gamma | 37, 39 |
| *Health care cost per overdose* |  |  |  |
| Permanent housing | 3,160 (1,353–6,303) | Gamma | 20 |
| Temporarily sheltered | 3,160 (1,353–6,303) | Gamma | 20, 38 |
| Unsheltered | 3,103 (1,329–6,191) | Gamma | 20, 38 |
| *Annual housing costs* |  |  |  |
| Permanent housing | 25,272 (16,355–36,098) | Gamma | 35 |
| Emergency shelter | 18,764 (12,143–26,802) | Gamma | 35 |
| Transitional shelter | 36,339 (23,517–51,907) | Gamma | 35 |
| **Quality-of-life Multipliers** |  |  |  |
| *OUD, in treatment* |  |  |  |
| Permanent housing | 0.721 (0.700–0.742) | Beta | 20 |
| Temporarily sheltered | 0.540 (0.524–0.555) | Beta | 20, 44 |
| Unsheltered | 0.244 (0.237–0.251) | Beta | 20, 44 |
| ***OUD, out of treatment*** |  |  |  |
| Permanent housing | 0.668 (0.655–0.680) | Beta | 20 |
| Temporarily sheltered | 0.500 (0.490–0.509) | Beta | 20, 44 |
| Unsheltered | 0.226 (0.221–0.230) | Beta | 20, 44 |
| ***OUD, abstinent<sup>d</sup>*** |  |  |  |
| Permanent housing |  |  |  |
| First month | 0.747 (0.728–0.766) | Beta | 20, 59 |
| Year $\geq 10$ | 0.983 (0.970–0.997) | Beta | 20, 59 |
| Temporarily sheltered |  |  |  |
| First month | 0.559 (0.545–0.573) | Beta | 20, 44, 59 |
| Year $\geq 10$ | 0.736 (0.726–0.746) | Beta | 20, 44, 59 |
| Unsheltered |  |  |  |
| First month | 0.253 (0.246–0.259) | Beta | 20, 44, 59 |
| Year $\geq 10$ | 0.332 (0.328–0.337) | Beta | 20, 44, 59 |

<sup>a</sup> Rates were derived from gamma-distributed all-cause and overdose mortality inputs; see Appendix C for the derivation.
<sup>b</sup> Rates of relapse from abstinence from second month to year 10 of abstinence were linear interpolations of the first month and year $\geq 10$ values.
<sup>c</sup> Rates were derived from beta-distributed annual probabilities; see Appendix D for the derivation.
<sup>d</sup> Utility values for second month through year 10 were linear interpolations of the first month and year $\geq 10$ values.

We adopted a lifetime horizon to evaluate long-term effects of the housing intervention. We used the model to calculate time spent in permanent housing, lifetime fatal overdoses, and mean age at death, as well as lifetime costs (2025 USD) and quality-adjusted life years (QALYs) for the cohort. We took a limited societal perspective that included healthcare and housing costs. We followed standard health economic modeling practices recommended by the Second Panel on Cost-Effectiveness in Health and Medicine, and discounted costs and QALYs at 3% per year.^21^ A Consolidated Health Economic Evaluation Reporting Standards (CHEERS) checklist and impact inventory are provided in **Appendices A and B**, respectively.^22^

### Housing and OUD states

We considered three housing states and three opioid use states (**Figure 1**). Each simulated individual was categorized by their housing status as either unsheltered (living in spaces not meant for habitation such as outdoors or in vehicles), sheltered but not stably housed (staying in emergency shelters or transitional shelters), or stably housed. Opioid use status was classified as opioid use without treatment (individuals not receiving medication-assisted treatment [MAT] for OUD), opioid use in treatment (receiving MAT), or abstinent (individuals who formerly used opioids and are not currently using opioids or receiving MAT for OUD).

**Figure 1.**
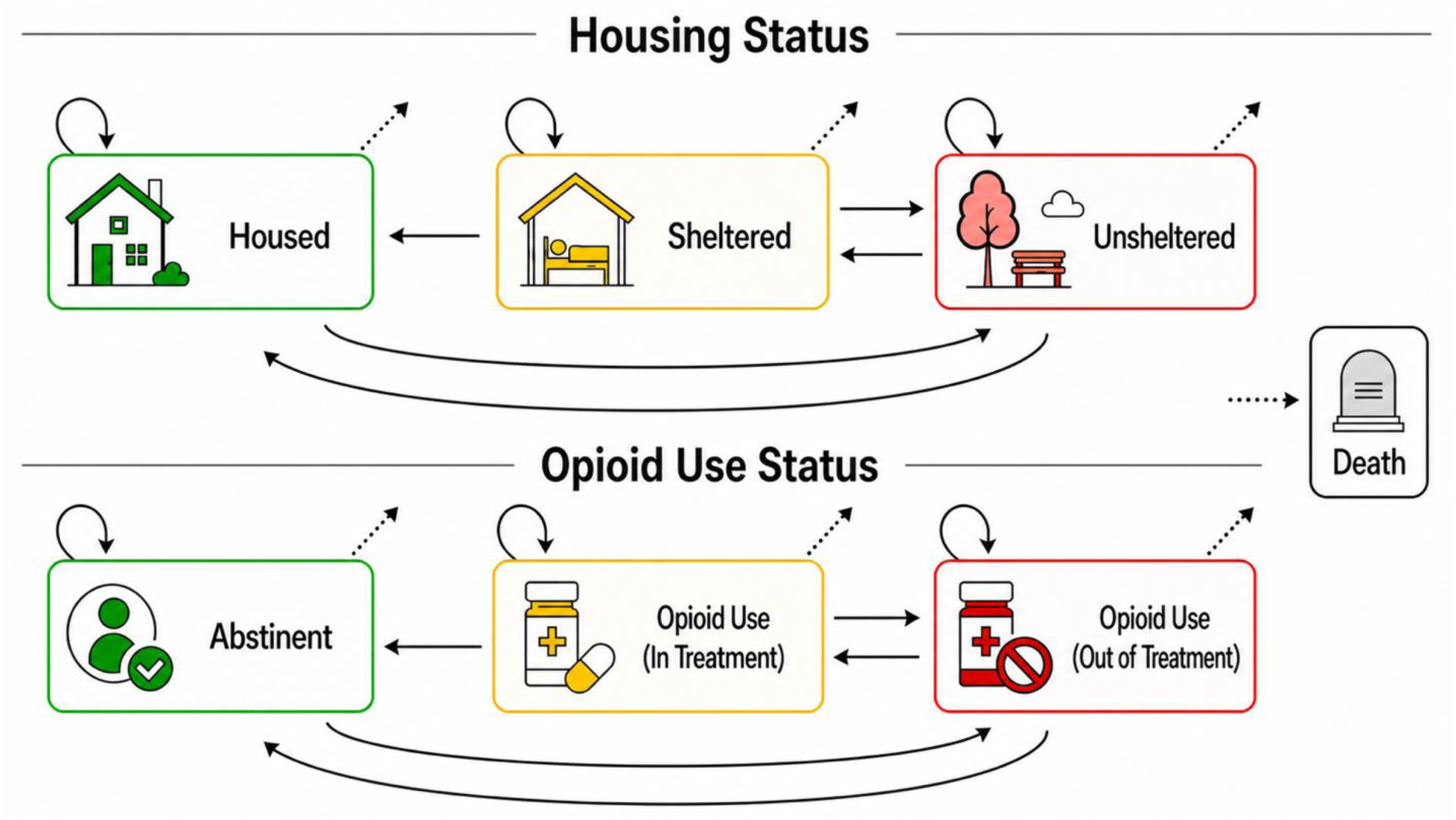
Model diagrams.

We allowed for all possible transitions between housing and opioid use states, with two exceptions. We assumed that abstinent individuals did not directly transition to OUD treatment when they relapsed but were instead initially out of treatment. We also assumed that permanently housed individuals did not directly transition to sheltered homelessness but were instead initially unsheltered when they lost access to housing.

### Death and Overdose

Mortality combined age-specific background risk from the 2021 United States life tables,^23^ homelessness-associated mortality multipliers,^24^ non-overdose excess mortality related to OUD, and fatal overdose. OUD-related mortality components were derived from a meta-analysis that separated overdose mortality from other causes of death.^25^ Overdose incidence varied by treatment status.^20^ Housing-specific overdose survival probabilities were derived using Coffin and Sullivan’s rescue model,^26^ data on fatal opioid overdoses stratified by homelessness status and documented other-person and naloxone presence,^27^ and evidence comparing fatal drug toxicity risk in public building versus outdoor settings (which informed the relative overdose fatality risk for sheltered versus unsheltered locations).^28^ Detailed derivations and assumptions are provided in **Appendix C**.

### OUD Transitions

Transition probabilities between OUD states (conditional on housed vs. unhoused) were calculated based on rates estimated in a prior study.^20^ Among PEH, only the probability of entering treatment differed between temporarily sheltered and unsheltered individuals, which was informed by a representative survey of adult PEH in California showing that unsheltered status was independently associated with worse health care access.^12^ All other OUD transition probabilities were assumed equal across the two unhoused states.

### Housing Transitions and Intervention

Housing transitions under the status quo were calibrated to Los Angeles County service data, including fiscal year 2023–24 measures of shelter access and permanent housing placement, interim housing exits, and returns to homelessness.^29–32^

We assumed a Housing First approach where PEH can acquire housing with no requirement to enter OUD treatment or achieve sobriety.^33^ We assumed that housing included standard supportive services associated with Housing First, such as intensive case management and access to and coordination with OUD treatment and educational, employment, mental health, and other social services.^14^ Among individuals initially unsheltered, the one-year access targets were 34.08% for temporary shelter and 8.87% for permanent housing. Following Measure A goals,^34^ the housing intervention increased these targets to 45.00% and 15.97%, respectively, while leaving other housing transition rates unchanged.^29^ Detailed derivations, calibration procedures, and internal validation are provided in **Appendix D**.

### Health and Economic Outcomes

Outcomes included permanent housing duration, fatal overdoses, age at death, discounted lifetime costs, and discounted QALYs. We calculated incremental cost-effectiveness ratios (ICERs) and incremental net monetary benefit (NMB).

Costs included healthcare and housing costs. Housing costs were adapted from published estimates.^35^ Temporary shelter costs were computed as a weighted average of emergency and transitional shelter costs (based on the observed prevalence of each shelter type). Health care costs combined age-and sex-specific background healthcare costs,^36^ with additional excess costs associated with OUD and homelessness, as well as OUD treatment and overdose costs, based on prior studies.^37, 38^ Medication costs used an adopted methadone and buprenorphine composite informed by treatment-utilization and cost sources.^39^

All costs were expressed in 2025 United States (U.S.) dollars. Healthcare costs were adjusted using the Personal Health Care expenditure price index with a Personal Consumption Expenditures healthcare bridge; housing costs were adjusted using the Personal Consumption Expenditures index.^40–42^ Health utility combined age-and sex-specific U.S. population values^43^ with housing-and OUD-specific multipliers,^20^ including a homelessness utility estimate of 0.43 from a national survey.^44^ We used differences in the prevalence of complex behavioral health needs as a proxy for differences in quality of life between sheltered and unsheltered individuals, assuming multipliers were inversely proportional to prevalence.^8^

### Statistical Analysis

All analyses were implemented in Python (version 3.12.2). Probabilistic sensitivity analysis (PSA) used 5,000 parameter set draws, with 50 stochastic replications per draw and strategy. Strategy-specific means across stochastic replications were paired within parameter draws, and 95% uncertainty intervals (UIs) were defined by the 2.5^th^ and 97.5^th^ percentiles across parameter draws. Convergence was assessed across parameter draws and stochastic replications, with details provided in **Appendix E**. Sensitivity analyses examined doubled permanent-housing costs, higher utility while unhoused (0.60 versus 0.43), and 20%, 40%, 60%, 80%, and 100% achievement of the targeted improvements in the housing intervention.

Our model directly generated population-level outcomes for California from the simulated cohort of 18,928 PEH with OUD. To estimate corresponding outcomes for Los Angeles County (which provided the primary empirical basis for the housing transition calibration and intervention targets), we scaled paired per-person incremental costs and QALYs to the estimated county population of PEH with OUD. The 2025 point-in-time count identified 65,584 adults aged 18 years or older experiencing homelessness in Los Angeles County.^45, 46^ Applying the same proxy for OUD (10.4% regular opioid use prevalence) yielded an estimated target population of 6,821 adults. Paired per-person incremental costs and QALYs were multiplied by the corresponding population size, assuming full implementation of the modeled housing intervention and unchanged per-person effects.

## RESULTS

### Base Case Analyses

The housing intervention increased mean permanent housing duration from 9.9 years (95% UI: 7.9–11.9) to 11.4 years (9.3–13.8). Mean age at death was 67.3 years (63.7–71.3) under the status quo and 67.6 years (64.0–71.6) with the housing intervention. Lifetime fatal overdoses in the cohort were 1,834 (946–3,163) and 1,795 (920–3,107), respectively. Discounted QALYs increased from 6.3 (95% UI: 5.4–7.2) to 6.8 (5.8–7.8) per person. Healthcare costs decreased from $336,557 ($289,157–$387,363) to $321,764 ($276,906–$370,028), while housing costs increased from $256,444 ($185,056–$339,324) to $287,300 ($206,379–$380,711).

The housing intervention generated 0.52 additional QALYs (95% UI: 0.45–0.61) at an incremental lifetime cost of $16,063 ($6,165-$27,400) per person, yielding an ICER of $31,083 ($11,865–$53,810) per QALY gained (**Figure 2**). At a willingness-to-pay threshold of $100K per QALY gained, incremental NMB was $35,615 ($22,773-$49,148).

**Figure 2.**
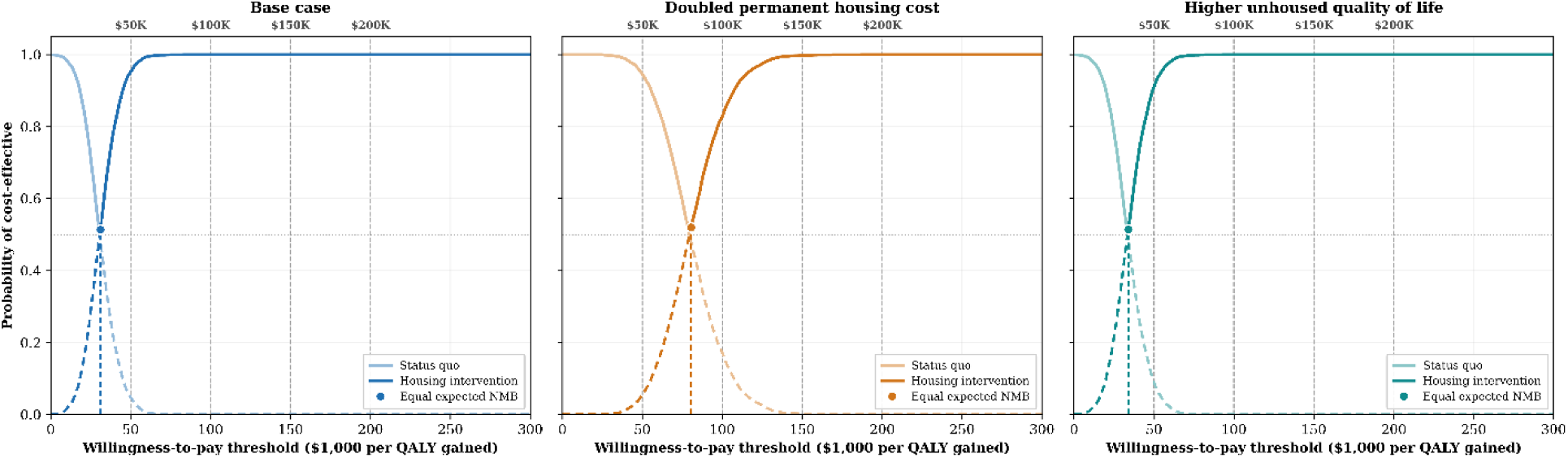
Cost-effectiveness acceptability frontiers (CEAF). The solid lines represent the CEAF, showing the probability that the strategy with the highest expected net monetary benefit is cost-effective at each willingness-to-pay threshold. The dashed lines show the corresponding cost-effectiveness acceptability curves for the status quo and housing intervention. The marked point identifies the willingness-to-pay threshold at which the status quo and housing intervention have equal expected net monetary benefit (NMB). QALY = quality-adjusted life year.

### Sensitivity Analysis

Doubling permanent housing costs increased incremental cost to $41,612 (95% UI: $22,791– $63,798) per person, with unchanged QALY gains, producing an ICER of $80,521 ($44,009– $124,750) per QALY gained; at a willingness-to-pay threshold of $100K per QALY gained, incremental NMB was $10,066 (-$12,555–$30,022). Higher utility while unhoused reduced incremental QALYs to 0.47 (0.41–0.57), with unchanged incremental costs, producing an ICER of $33,999 ($12,941–$58,785) per QALY gained; at a willingness-to-pay threshold of $100K per QALY gained, incremental NMB was $31,182 ($18,509-$44,654). Detailed scenario outcomes are reported in **Table 2** and **Appendix F**.

**Table 2.**
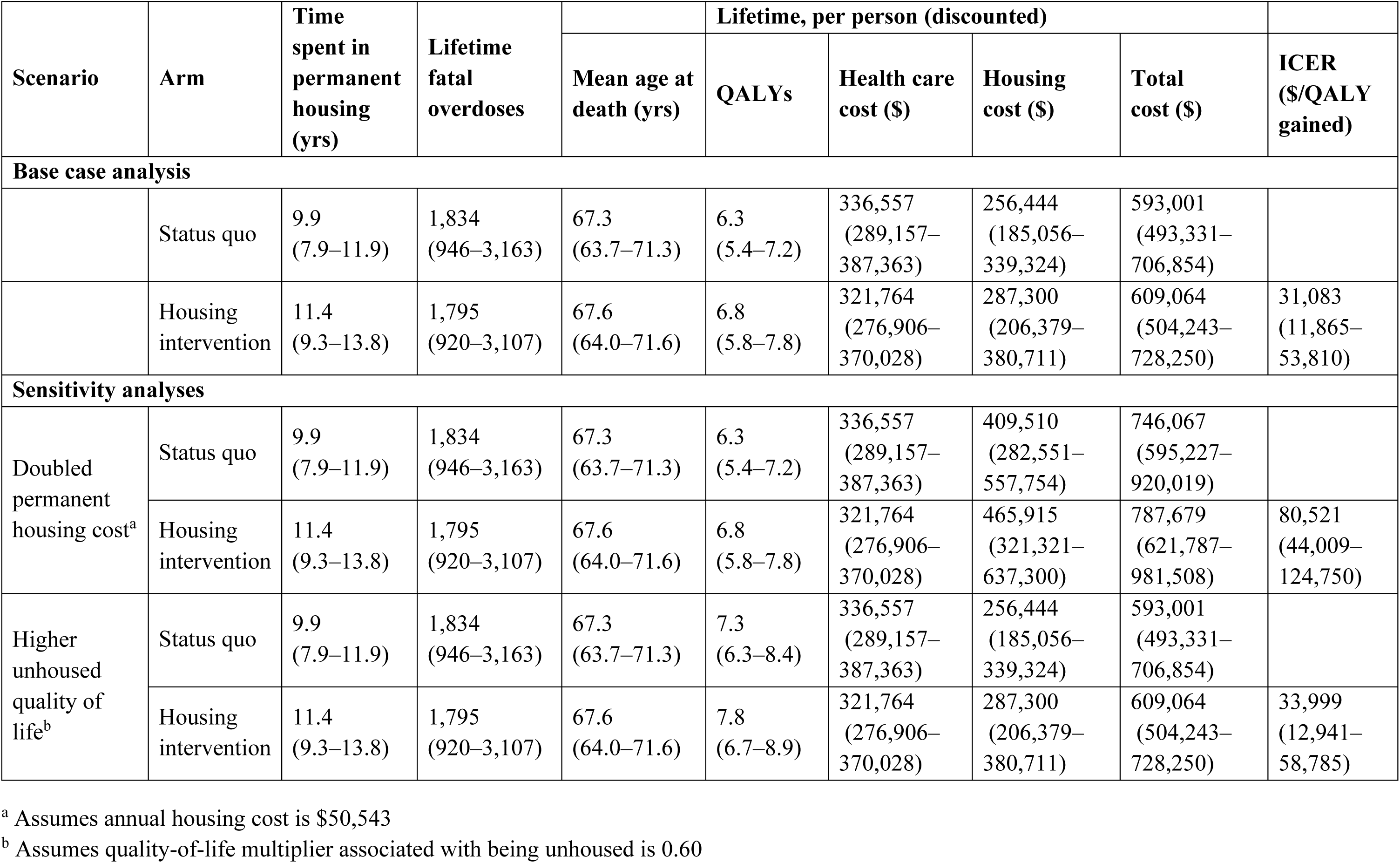
Outcomes for Each Scenario, Mean and 95% Uncertainty Intervals.

| Scenario | Arm | Time spent in permanent housing (yrs) | Lifetime fatal overdoses | | Lifetime, per person (discounted) | | | | ICER (\$/QALY gained) |
| --- | --- | --- | --- | --- | --- | --- | --- | --- | --- |
| | | | | Mean age at death (yrs) | QALYs | Health care cost (\$) | Housing cost (\$) | Total cost (\$) | |
| Base case analysis |  |  |  |  |  |  |  |  |  |
|  | Status quo | 9.9<br>(7.9–11.9) | 1,834<br>(946–3,163) | 67.3<br>(63.7–71.3) | 6.3<br>(5.4–7.2) | 336,557<br>(289,157–387,363) | 256,444<br>(185,056–339,324) | 593,001<br>(493,331–706,854) |  |
|  | Housing intervention | 11.4<br>(9.3–13.8) | 1,795<br>(920–3,107) | 67.6<br>(64.0–71.6) | 6.8<br>(5.8–7.8) | 321,764<br>(276,906–370,028) | 287,300<br>(206,379–380,711) | 609,064<br>(504,243–728,250) | 31,083<br>(11,865–53,810) |
| Sensitivity analyses |  |  |  |  |  |  |  |  |  |
| Doubled permanent housing cost<sup>a</sup> | Status quo | 9.9<br>(7.9–11.9) | 1,834<br>(946–3,163) | 67.3<br>(63.7–71.3) | 6.3<br>(5.4–7.2) | 336,557<br>(289,157–387,363) | 409,510<br>(282,551–557,754) | 746,067<br>(595,227–920,019) |  |
|  | Housing intervention | 11.4<br>(9.3–13.8) | 1,795<br>(920–3,107) | 67.6<br>(64.0–71.6) | 6.8<br>(5.8–7.8) | 321,764<br>(276,906–370,028) | 465,915<br>(321,321–637,300) | 787,679<br>(621,787–981,508) | 80,521<br>(44,009–124,750) |
| Higher unhoused quality of life<sup>b</sup> | Status quo | 9.9<br>(7.9–11.9) | 1,834<br>(946–3,163) | 67.3<br>(63.7–71.3) | 7.3<br>(6.3–8.4) | 336,557<br>(289,157–387,363) | 256,444<br>(185,056–339,324) | 593,001<br>(493,331–706,854) |  |
|  | Housing intervention | 11.4<br>(9.3–13.8) | 1,795<br>(920–3,107) | 67.6<br>(64.0–71.6) | 7.8<br>(6.7–8.9) | 321,764<br>(276,906–370,028) | 287,300<br>(206,379–380,711) | 609,064<br>(504,243–728,250) | 33,999<br>(12,941–58,785) |

<sup>a</sup> Assumes annual housing cost is \$50,543
<sup>b</sup> Assumes quality-of-life multiplier associated with being unhoused is 0.60

No break-even effectiveness level was identified between 20% and 100% achievement of the housing intervention. At 20% achievement of the housing intervention, at a willingness-to-pay threshold of $100K per QALY gained, incremental NMB was $8,414 (95% UI: $5,307–$11,970) under the base case and $2,339 (-$3,044–$7,181) when permanent housing costs were doubled.

### Population-Level Implications

For the estimated 18,928 PEH with OUD in California, the housing intervention was projected to generate 9,782 (95% UI: 8,427–11,551) additional discounted QALYs and $304.0 (95% UI: $116.7–$518.6) million in additional discounted lifetime costs. The additional lifetime cost reflected approximately $584.0 million in higher housing expenditures, partly offset by $280.0 million in health care savings. For the estimated Los Angeles County target population of 6,821 adults, the projected gains were 3,525 (95% UI: 3,037–4,162) QALYs with $110 (95% UI: $42– $187) million in additional discounted lifetime costs, reflecting approximately $211 million in higher housing expenditures offset by $101 million in health care savings.

## DISCUSSION

In this model-based economic evaluation, increasing permanent housing access for PEH with OUD in California improved health outcomes and was cost-effective compared to the alternative of not increasing housing access. In the base case, the housing intervention produced an ICER of $31,083 per QALY gained, a value below commonly used U.S. willingness-to-pay thresholds. These findings are consistent with prior modeling work showing that stable housing for PEH with OUD can provide health gains at an ICER near $26,800 per QALY gained.^20^ Our analysis extended this work by representing housing as a dynamic process rather than a one-time transition into stable housing, and by distinguishing unsheltered from temporarily sheltered states. In the context of federal shifts away from Housing First approaches, our policy analysis contributes evidence on how investments in stable housing, without requirements of OUD treatment, impact short-and long-term health outcomes, spending on housing, and spending on healthcare.

Housing should be understood as a structural component of OUD care rather than a substitute for addiction treatment. A randomized trial demonstrated that a Housing First approach could achieve earlier and more stable housing without requiring abstinence.^47^ In the At Home/Chez Soi project, rent supplements and intensive case management improved housing stability, although improvements in generic quality of life were less consistent, and extended follow-up did not identify an overall mortality difference.^48, 49^ At the same time, Housing First did not significantly improve methadone adherence in a randomized trial subgroup.^50^ Conversely, receipt of methadone or buprenorphine after nonfatal overdose has been associated with lower mortality,^51^ and low-barrier buprenorphine programs have demonstrated that people experiencing homelessness can be engaged in treatment without first achieving housing stability.^52^ Therefore, this evidence supports treating housing, OUD treatment, and overdose prevention as complementary services. Housing access need not be conditioned on treatment participation or abstinence, while treatment and overdose prevention services should remain readily accessible before and after housing placement.

The reduction in healthcare spending projected in our analysis is also consistent with evidence that housing interventions can reduce or shift use of health services, although the magnitude of these effects varies across settings. Housing and case management reduced acute care use among chronically ill adults experiencing homelessness in Chicago.^53^ A Santa Clara County, California trial found reductions in psychiatric emergency visits and shelter use but no significant reductions in overall emergency visits or hospitalizations,^54^ while a Denver Housing First evaluation found greater use of psychiatric outpatient care and prescriptions alongside fewer emergency visits.^55^ Economic evaluations of At Home/Chez Soi similarly found that reductions in other service use offset part of housing program expenditures.^56, 57^ Although these studies differ in population, intervention, setting, and follow-up, they reinforce the importance of considering both health benefits and changes in resource use when evaluating housing interventions rather than expecting health care savings alone to finance housing programs.

Our sensitivity analyses highlighted the importance of local housing costs and the extent to which improvements in housing access can be achieved. Higher permanent housing costs had a greater influence on economic outcomes than the alternative assumption about health utility while unhoused. Importantly, mean net benefit remained favorable across all evaluated levels of achievement of the housing access targets, including when only a portion of the targeted improvement was realized. The population-level findings further suggested that expanding housing access provides good value, while still requiring substantial spending to implement at scale.^58^ Housing expenditures and health care savings may accrue to different agencies and at different times, creating financing challenges even when an intervention provides favorable long-term value. Budget planning therefore requires consideration of implementation pace, available housing capacity, population turnover, service capacity, and the timing and distribution of expenditures across sectors. These considerations are particularly relevant for California policymakers allocating resources across homelessness services, addiction treatment, and overdose prevention.

Several limitations merit consideration. First, model inputs were drawn from observational studies, administrative records, and prior models, which may introduce uncertainty when combining evidence from different sources. Second, housing-specific costs and utilities were applied when individuals changed housing states, although observed differences between housing groups may partly reflect pre-existing health differences rather than effects of housing. Third, housing transition parameters were informed by Los Angeles County data and may not fully represent other California jurisdictions or settings with different service systems, housing markets, and implementation environments. Fourth, our limited societal perspective did not include potential criminal justice cost savings associated with transitions to permanent housing. To the extent that such savings occur, their inclusion would make the housing intervention appear more economically favorable. Finally, as with any model-based analysis, the results depend on structural assumptions that simplify complex real-world processes. These limitations introduce uncertainty around the magnitude of the projected benefits. Nonetheless, our findings suggest that expanded housing access could improve health outcomes for PEH with OUD.

## CONCLUSIONS

Expanded housing access for PEH with OUD should be considered not only as social policy but also as health policy. In our model, a Housing First style intervention increased the time spent in permanent housing, improved quality-adjusted survival, and reduced fatal overdoses. For California decision makers, our findings support considering housing alongside addiction treatment and overdose prevention when setting priorities, allocating resources, and planning budgets across housing and health systems. More broadly, coordinated cross-sector investment may help align resources devoted to homelessness and OUD with interventions that generate both housing and health benefits.

## Acknowledgments

This work was supported by Grant R37-DA15612 and Grant R01-DA064386 from the National Institute on Drug Abuse. The funders had no role in study design, data collection and analysis, decision to publish, or preparation of the manuscript.

## AI Disclosure

We used ChatGPT (OpenAI; GPT-5.5) to write some sections of the code. Two authors independently reviewed, tested, and validated the AI-generated code.

## Data Availability

All data produced in the present work are contained in the manuscript. The analytic code is publicly accessible at https://github.com/yuewei-ling/PEH-OUD_Microsimulation.

