## Supplementary Materials for "Cost-Effectiveness of Expanding Housing Access for People Experiencing Homelessness and Opioid Use Disorder in California"

**Appendix A. CHEERS Checklist**

| **Topic** | **No.** | **Item** | **Location where item is reported** |
|---|---|---|---|
| **Title** | | | |
| | 1 | Identify the study as an economic evaluation and specify the interventions being compared. | Title |
| **Abstract** | | | |
| | 2 | Provide a structured summary that highlights context, key methods, results, and alternative analyses. | Abstract |
| **Introduction** | | | |
| **Background and objectives** | 3 | Give the context for the study, the study question, and its practical relevance for decision making in policy or practice. | Introduction |
| **Methods** | | | |
| **Health economic analysis plan** | 4 | Indicate whether a health economic analysis plan was developed and where available. | No pre-registered health economic analysis plan exists for this analysis. |
| **Study population** | 5 | Describe characteristics of the study population (such as age range, demographics, socioeconomic, or clinical characteristics). | Methods ("Overview");<br>Table 1 |
| **Setting and location** | 6 | Provide relevant contextual information that may influence findings. | Methods ("Overview", "Housing Transitions and Intervention");<br>Appendix D |
| **Comparators** | 7 | Describe the interventions or strategies being compared and why chosen. | Methods ("Housing Transitions and Intervention") |
| **Perspective** | 8 | State the perspective(s) adopted by the study and why chosen. | Methods ("Overview");<br>Impact Inventory |

| | | | |
|---|---|---|---|
| **Time horizon** | 9 | State the time horizon for the study and why appropriate. | Methods ("Overview") |
| **Discount rate** | 10 | Report the discount rate(s) and reason chosen. | Methods ("Overview") |
| **Selection of outcomes** | 11 | Describe what outcomes were used as the measure(s) of benefit(s) and harm(s). | Methods ("Health and Economic Outcomes") |
| **Measurement of outcomes** | 12 | Describe how outcomes used to capture benefit(s) and harm(s) were measured. | Methods ("Death and Overdose", "Health and Economic Outcomes");<br>Appendix C;<br>Appendix D |
| **Valuation of outcomes** | 13 | Describe the population and methods used to measure and value outcomes. | Methods ("Health and Economic Outcomes");<br>Table 1 |
| **Measurement and valuation of resources and costs** | 14 | Describe how costs were valued. | Methods ("Health and Economic Outcomes");<br>Table 1 |
| **Currency, price date, and conversion** | 15 | Report the dates of the estimated resource quantities and unit costs, plus the currency and year of conversion. | Methods ("Health and Economic Outcomes");<br>Table 1 |
| **Rationale and description of model** | 16 | If modelling is used, describe in detail and why used. Report if the model is publicly available and where it can be accessed. | Methods ("Overview", "Housing and OUD States", "OUD Transitions", "Housing Transitions and Intervention");<br>Figure 1 |
| **Analytics and assumptions** | 17 | Describe any methods for analysing or statistically transforming data, any extrapolation methods, and approaches for validating any model used. | Methods ("Housing and OUD states", "Housing Transitions and Intervention", "Health and Economic Outcomes", "Statistical Analysis");<br>Appendix C;<br>Appendix D;<br>Appendix E |
| **Characterising heterogeneity** | 18 | Describe any methods used for estimating how the results of the study vary for subgroups. | N/A |

| | | | |
|---|---|---|---|
| **Characterising distributional effects** | 19 | Describe how impacts are distributed across different individuals or adjustments made to reflect priority populations. | N/A |
| **Characterising uncertainty** | 20 | Describe methods to characterise any sources of uncertainty in the analysis. | Methods ("Statistical Analysis"); Table 1 |
| **Approach to engagement with patients and others affected by the study** | 21 | Describe any approaches to engage patients or service recipients, the general public, communities, or stakeholders (such as clinicians or payers) in the design of the study. | N/A |
| **Results** | | | |
| **Study parameters** | 22 | Report all analytic inputs (such as values, ranges, references) including uncertainty or distributional assumptions. | Table 1; Appendix C; Appendix D |
| **Summary of main results** | 23 | Report the mean values for the main categories of costs and outcomes of interest and summarise them in the most appropriate overall measure. | Results ("Base Case Analyses"); Figure 2; Table 2; Appendix F |
| **Effect of uncertainty** | 24 | Describe how uncertainty about analytic judgments, inputs, or projections affect findings. Report the effect of choice of discount rate and time horizon, if applicable. | Results ("Sensitivity Analysis"); Figure 2; Table 2; Appendix F |
| **Effect of engagement with patients and others affected by the study** | 25 | Report on any difference patient/service recipient, general public, community, or stakeholder involvement made to the approach or findings of the study | N/A |
| **Discussion** | | | |
| **Study findings, limitations,** | 26 | Report key findings, limitations, ethical or equity | Discussion; Limitations |

| | | | |
|---|---|---|---|
| **generalisability, and current knowledge** | | considerations not captured, and how these could affect patients, policy, or practice. | |
| **Other relevant information** | | | |
| **Source of funding** | 27 | Describe how the study was funded and any role of the funder in the identification, design, conduct, and reporting of the analysis | Title page (“Acknowledgments”) |
| **Conflicts of interest** | 28 | Report authors conflicts of interest according to journal or International Committee of Medical Journal Editors requirements. | Title page (“Declarations”) |

### Appendix B. Impact Inventory

| Sector | Type of Impact | Case Analysis Perspective<br>Limited Societal | Notes on Sources of Evidence |
|---|---|---|---|
| **Formal Healthcare Sector** | | | |
| **Health** | **Health outcomes (effects)** | | |
| | Longevity effects | ✓ | Health outcomes are derived from the literature; see Table 1, Methods ("Health and Economic Outcomes"), and Appendix C. |
| | Health-related quality-of-life effects | ✓ | |
| | Other health effects | ✓ | |
| | **Medical costs** | | |
| | Paid for by third-party payers | ✓ | Medical costs are derived from the literature; see Table 1 and Methods ("Health and Economic Outcomes"). |
| | Paid for by patients out-of-pocket | ✓ | |
| | Future related medical costs (payers and patients) | ✓ | |
| | Future unrelated medical costs (payers and patients) | ✓ | |
| **Informal Healthcare Sector** | | | |
| **Health** | Patient-time costs | ✕ | |
| | Unpaid caregiver-time costs | ✕ | |
| | Transportation costs | ✕ | |
| **Non-Healthcare Sectors (with examples of possible items)** | | | |
| **Productivity** | Labor-market earnings lost | ✕ | |
| | Cost of unpaid lost productivity due to illness | ✕ | |
| | Cost of uncompensated household productivity | ✕ | |

| | | | |
|---|---|---|---|
| **Consumption** | Future consumption unrelated to health | ✕ | |
| **Social Services** | Cost of social services as part of the intervention | ✓ | Housing costs are derived from published estimates of the housing continuum; see Table 1 and Methods ("Health and Economic Outcomes"). |
| **Legal or Criminal Justice** | Number of crimes related to the intervention | ✕ | |
| | Cost of crimes related to the intervention | ✕ | |
| **Education** | Impact on educational achievement of the population | NA | |
| **Housing** | Cost of the intervention on home improvements | NA | |
| **Environment** | Production of toxic-waste pollution | NA | |
| **Other** | Other impacts | NA | |

### Appendix C. Mortality and Overdose

For each living individual, the annual mortality hazard combined age-specific background mortality multiplied by the housing-related mortality multiplier, non-overdose excess mortality related to opioid use disorder (OUD), and fatal overdose:

$$\lambda_{\text{death}}(a, h, o) = M(a, h) \cdot \lambda_{\text{background}}(a) + \lambda_{\text{OUD,non-overdose}}(o) + \lambda_{\text{fatal\_overdose}}(h, o) \tag{C1}$$

where $a$ denotes age, $h$ housing state, $o$ OUD state, $M$ the homeless multiplier, and $\lambda$ an annual hazard.

#### *Non-overdose excess mortality*

Ma et al. reported pooled crude mortality rates among people with OUD during medication treatment, untreated periods, and after treatment cessation.[C1] The all-cause mortality rates were 0.92 (95% CI, 0.79–1.04), 4.89 (3.54–6.23), and 1.69 (1.47–1.91) per 100 person-years, respectively. The corresponding overdose mortality rates were 0.24 (0.20–0.28), 2.43 (1.72–3.15), and 0.68 (0.55–0.80) per 100 person-years, respectively. The post-treatment-cessation mortality estimate was used as a proxy for the abstinent state. We used a background mortality rate of 0.0028 per person-year, corresponding to the US median age of 39.2 years based on CDC life tables, as the background mortality anchor.[C2,C3] The age-independent OUD-related non-overdose excess mortality hazard was calculated by subtracting both the overdose mortality component and the background-mortality anchor from the corresponding all-cause mortality rate:

$$\lambda_{\text{OUD,non-overdose}}^{\text{in treatment}} = 0.0092 - 0.0024 - 0.0028 = 0.0040/\text{year} \tag{C2}$$

$$\lambda_{\text{OUD,non-overdose}}^{\text{out of treatment}} = 0.0489 - 0.0243 - 0.0028 = 0.0218/\text{year} \tag{C3}$$

$$\lambda_{\text{OUD,non-overdose}}^{\text{abstinent}} = 0.0169 - 0.0068 - 0.0028 = 0.0073/\text{year} \tag{C4}$$

#### *Housing-specific overdose survival probability*

We defined three mutually exclusive record categories: $C_0$, other-person presence not recorded; $C_1$, other-person presence recorded but naloxone not recorded within that subgroup; and $C_2$, other-person presence and naloxone both recorded. These categories described documentation rather than confirmed absence of rescue, timely witnessing, or adequate treatment.

Coffin and Sullivan reported model-summary survival probabilities of 0.899 without medical assistance or lay naloxone and 0.978 with emergency medical services or naloxone assistance.[C4] We therefore assigned:

$$s_0 = 0.899 \tag{C5}$$

$$s_2 = 0.978 \tag{C6}$$

For $C_1$ (other-person presence recorded but naloxone not recorded), absence of a naloxone record was not assumed to exclude other medical assistance. We set the probability of other modeled rescue to 0.40, adapting Coffin and Sullivan's emergency medical services call probability for subsequent overdoses.[C4] The resulting survival probability was:

$$s_1 = 0.60(0.899) + 0.40(0.978) = 0.9306 \tag{C7}$$

Booth et al. reported 5,760 opioid-related overdose deaths among individuals not identified as experiencing homelessness and 884 among individuals identified as experiencing homelessness in Ontario.[C5] These groups were used as proxies for housed population and unhoused (combined temporarily sheltered and unsheltered) population, respectively. Among these deaths, 2,862 and 493 decedents, respectively, were recorded as not alone at the time of death. Naloxone administration was recorded for 828 and 203 of these decedents. These data yielded 2,898, 2,034, and 828 deaths across $C_0$, $C_1$, and $C_2$ for the housed proxy group, and 391, 290, and 203 deaths, respectively, for the unhoused proxy group.

For each record category, the model-implied number of overdose episodes was calculated by dividing the observed number of deaths by the corresponding probability of death. Therefore, the overdose survival probabilities were:

$$\begin{aligned} s_{\text{housed}} &= 1 - \left( \sum_{k=0}^{2} \frac{d_{k,\text{housed}} / \sum_{k=0}^{2} d_{k,\text{housed}}}{1 - s_k} \right)^{-1} \\ &= 1 - \frac{5760}{\frac{2898}{(1-0.899)} + \frac{2034}{(1-0.9306)} + \frac{828}{(1-0.978)}} \\ &= 0.9397727616 \end{aligned} \tag{C8}$$

$$\begin{aligned} s_{\text{unhoused}} &= 1 - \left( \sum_{k=0}^{2} \frac{d_{k,\text{unhoused}} / \sum_{k=0}^{2} d_{k,\text{unhoused}}}{1 - s_k} \right)^{-1} \\ &= 1 - \frac{884}{\frac{391}{(1-0.899)} + \frac{290}{(1-0.9306)} + \frac{203}{(1-0.978)}} \\ &= 0.9488344032 \end{aligned} \tag{C9}$$

Hu et al. reported fully adjusted odds ratios for fatal drug toxicity in public buildings and outdoor settings, relative to private locations, of 0.08 and 0.16, respectively.[C6] We used their ratio, $0.08/0.16 = 0.50$, as a proxy for the sheltered-to-unsheltered contrast in the odds of fatal overdose:

$$\frac{(1 - s_{\text{sheltered}})/s_{\text{sheltered}}}{(1 - s_{\text{unsheltered}})/s_{\text{unsheltered}}} = 0.50 \tag{C10}$$

In our microsimulation, 22.4% of the initially unhoused population was temporarily sheltered and 77.6% was unsheltered. We used these proportions as overdose-event weights, assuming comparable average overdose incidence across the two groups in the reference population. The weighted survival probability was constrained to equal the previously derived survival probability for the combined unhoused proxy group:

$$0.224 s_{\text{sheltered}} + 0.776 s_{\text{unsheltered}} = 0.9488344032 \quad \text{(C11)}$$

Solving equations C10 and C11 yielded overdose survival probabilities of 0.970449 for temporarily sheltered individuals and 0.942595 for unsheltered individuals.

***References***

[C1] Ma J, Bao YP, Wang RJ, et al. Effects of medication-assisted treatment on mortality among opioids users: a systematic review and meta-analysis. *Mol Psychiatry*. 2019;24:1868-1883.

[C2] United States Census Bureau. American Community Survey. Accessed Oct 1, 2025. https://data.census.gov/all?q=american%20community%20survey

[C3] Arias E, Xu J, Kochanek K. United States life tables, 2021. *Natl Vital Stat Rep*. 2023;72(12).

[C4] Coffin PO, Sullivan SD. Cost-effectiveness of distributing naloxone to heroin users for lay overdose reversal. Ann Intern Med. 2013;158(1):1–9.

[C5] Booth RG, Shariff SZ, Carter B, Hwang SW, Orkin AM, Forchuk C, et al. Opioid-related overdose deaths among people experiencing homelessness, 2017 to 2021: a population-based analysis using coroner and health administrative data from Ontario, Canada. Addiction. 2024;119(2):334–344.

[C6] Hu K, Klinkenberg B, Gan WQ, Slaunwhite AK. Spatial-temporal trends in the risk of illicit drug toxicity death in British Columbia. BMC Public Health. 2022;22:2121.

### Appendix D. Housing Transitions and Intervention

#### *Housing Transitions*

Los Angeles County administrative data on homeless services were used to parameterize housing transitions and the housing intervention. In the absence of comparable statewide transition data for people experiencing homelessness (PEH), we assumed that Los Angeles County housing system dynamics provided a reasonable approximation for the modeled population (PEH with OUD in California).

For transitions out of temporary shelter, Los Angeles County reported 5,054 exits to permanent housing among 23,275 active interim housing participants, corresponding to an annual proportion of 0.217143.[D1] An earlier county evaluation reported 3,582 exits to unsheltered homelessness among 44,840 interim housing enrollees, corresponding to an annual proportion of 0.079884.[D2] The permanent housing and unsheltered homelessness represent competing exits from temporary shelter, so we first converted their combined annual probability to a continuous-time annual rate:

$$r_{\text{leaving temporary shelter}} = -\ln[1-(0.217143+0.079884)] = 0.352437/\text{year} \quad \text{(D1)}$$

We then allocated this rate between permanent housing and unsheltered homelessness in proportion to the corresponding annual exit proportions:

$$r_{\text{temporarily sheltered}\rightarrow\text{permanent housing}} = r_{\text{leaving temporary shelter}} * \left(\frac{0.217143}{0.217143+0.079884}\right) = 0.2577/\text{year} \quad \text{(D2)}$$

$$r_{\text{temporarily sheltered}\rightarrow\text{unsheltered}} = r_{\text{leaving temporary shelter}} * \left(\frac{0.079884}{0.217143+0.079884}\right) = 0.0948/\text{year} \quad \text{(D3)}$$

The Measure A Baselines Report estimated a 12-month return-to-homelessness probability of 13.4%.[D3] We used this as the annual probability of housing loss and converted it to a continuous-time rate:

$$r_{\text{permanent housing}\rightarrow\text{unsheltered}} = -\ln(1-0.134) = 0.1439/\text{year} \quad \text{(D4)}$$

The Measure A baseline materials reported 66,919 service participants who experienced unsheltered homelessness during fiscal year 2023–24, of whom 22,804 accessed interim housing.[D4] We therefore used $22{,}804/66{,}919 = 0.340770$ as the one-year status quo target for ever accessing temporary shelter among individuals initially unsheltered. The corresponding permanent housing target was based on 5,937 placements from unsheltered settings, yielding a one-year target of $5{,}937/66{,}919 = 0.088719$.[D5] The annual transition rates from unsheltered homelessness to temporary shelter and permanent housing were calibrated jointly to reproduce the two one-year access targets.

We developed a continuous-time housing model to track both current housing state and whether temporary shelter or permanent housing had previously been reached, since individuals could move repeatedly among unsheltered homelessness, temporary shelter, and permanent housing during the year. For each candidate pair of transition rates from unsheltered homelessness to temporary shelter and permanent housing, the model calculated the exact one-year probabilities of ever reaching each destination while holding the other housing transition rates fixed. The two rates were then solved jointly so that the model-predicted cumulative access probabilities matched the calibration targets. This yielded $r_{\text{unsheltered}\rightarrow\text{temporarily sheltered}} = 0.4302/\text{year}$ and $r_{\text{unsheltered}\rightarrow\text{permanent housing}} = 0.0584/\text{year}$.

#### *Housing Intervention*

The housing intervention is informed by Los Angeles County Measure A targets. Measure A established a goal of increasing interim housing access among people experiencing unsheltered homelessness from approximately 34% to 45%.[D5] We therefore set the one-year temporary-shelter access target to 45.00%. Measure A also increased the annual target for permanent-housing placements from unsheltered settings from 5,937 to 10,687.[D5] Holding the fiscal year 2023–24 unsheltered service participant denominator of 66,919 fixed gave a one-year permanent-housing access target of $10{,}687/66{,}919 = 0.159701$.

The housing intervention only changed the transition rates from unsheltered homelessness to temporary shelter and permanent housing. Applying the same joint calibration procedure to the intervention targets yielded $r^{*}_{\text{unsheltered}\rightarrow\text{temporary shelter}} = 0.6506/\text{year}$ and $r^{*}_{\text{unsheltered}\rightarrow\text{permanent housing}} = 0.1459/\text{year}$. All other housing transition rates remained unchanged.

To evaluate the effectiveness of the housing intervention, one-year access targets were interpolated between the status quo and housing intervention at 20%, 40%, 60%, and 80% of the targeted improvement. Each set of targets was calibrated using the same procedure.

#### *Internal Validation*

Housing transition rates were calibrated in the continuous-time housing model and subsequently implemented in the monthly microsimulation. We assessed the internal consistency by comparing the targeted one-year cumulative probabilities of accessing temporary shelter or permanent housing with the corresponding probabilities produced by the housing model and the microsimulation among individuals initially unsheltered.

Under the status quo, the probability of temporary shelter access was 33.41%, compared with the 34.08% target, and permanent housing access was 8.36%, compared with the 8.87% target.

Under the housing intervention, the corresponding probabilities were 44.14% vs. 45.00% for temporary shelter and 15.21% vs. 15.97% for permanent housing. Across all scenarios, the microsimulation estimates were close to the corresponding calibration targets (**Table D1**).

**Table D1. One-year housing access targets and model outcomes.**

| Transitions | Target (%) | Housing Model (%) | Microsimulation (%) |
|---|---|---|---|
| Status quo | | | |
| Unsheltered to temporarily sheltered | 34.079 | 34.069 | 33.410 |
| Unsheltered to permanent housing | 8.872 | 8.553 | 8.362 |
| 20% of the housing intervention | | | |
| Unsheltered to temporarily sheltered | 36.262 | 36.248 | 35.553 |
| Unsheltered to permanent housing | 10.292 | 9.951 | 9.731 |
| 40% of the housing intervention | | | |
| Unsheltered to temporarily sheltered | 38.446 | 38.429 | 37.698 |
| Unsheltered to permanent housing | 11.711 | 11.348 | 11.101 |
| 60% of the housing intervention | | | |
| Unsheltered to temporarily sheltered | 40.631 | 40.609 | 39.845 |
| Unsheltered to permanent housing | 13.131 | 12.744 | 12.471 |
| 80% of the housing intervention | | | |
| Unsheltered to temporarily sheltered | 42.815 | 42.789 | 41.993 |
| Unsheltered to permanent housing | 14.550 | 14.141 | 13.841 |
| Housing intervention | | | |
| Unsheltered to temporarily sheltered | 45.000 | 44.969 | 44.140 |
| Unsheltered to permanent housing | 15.970 | 15.537 | 15.211 |

***References***

[D1] Los Angeles County Homeless Initiative. Quarterly Report No. 30, FY2023–24 housing-program performance tables. December 10, 2024. Available at: https://file.lacounty.gov/SDSInter/lac/1174239_HomelessInitiativeQuarterlyReportNo.30-SIGNEDBOARDMEMO_1_.pdf

[D2] Toros H, Culhane D, Metraux S. Annual performance evaluation for year 6 of the Countywide Homeless Initiative. 2024.

[D3] Los Angeles County Measure A Data Subcommittee. Measure A Baselines Report. January 24, 2025. Available at: https://file.lacounty.gov/SDSInter/lac/1187058_DataSubcommitteeBaselinesreport.pdf

[D4] Los Angeles County. Measure A Baselines: Demographics Memo and Tables. Measure A Data Subcommittee / Los Angeles County Homeless Initiative. March 3, 2025. Available at: https://file.lacounty.gov/SDSInter/lac/1187077_Measure-A-Baselines-Demographics-memo-and-tables.pdf

[D5] Los Angeles County Homeless Initiative. Measure A Resource Hub: adopted goals, metrics, baselines, and 2030 targets. Available at: https://homeless.lacounty.gov/measure-a-hub/

### Appendix E. Simulation Convergence

We simulated a cohort of 18,928 PEH with OUD, using 5,000 probabilistic parameter draws and 50 stochastic replications per draw. We evaluated numerical convergence separately across probabilistic parameter draws and stochastic replications.

As the number of PSA draws increased, the running means of incremental costs, incremental quality-adjusted life years (QALYs), and incremental net monetary benefit (NMB) at a willingness-to-pay threshold of $100,000 per QALY gained became progressively more stable, while the Monte Carlo confidence bands narrowed (**Figure E1**). Incremental QALYs remained within 1% of the final mean from draw 61 onward, compared with draw 389 for INMB and draw 567 for incremental cost. At 5,000 draws, the relative Monte Carlo standard errors were 0.113% for incremental QALYs, 0.263% for INMB, and 0.472% for incremental cost.

**Figure E1. Convergence across parameter draws.** PSA = probabilistic sensitivity analysis, QALYs = quality-adjusted life years, NMB = net monetary benefit.

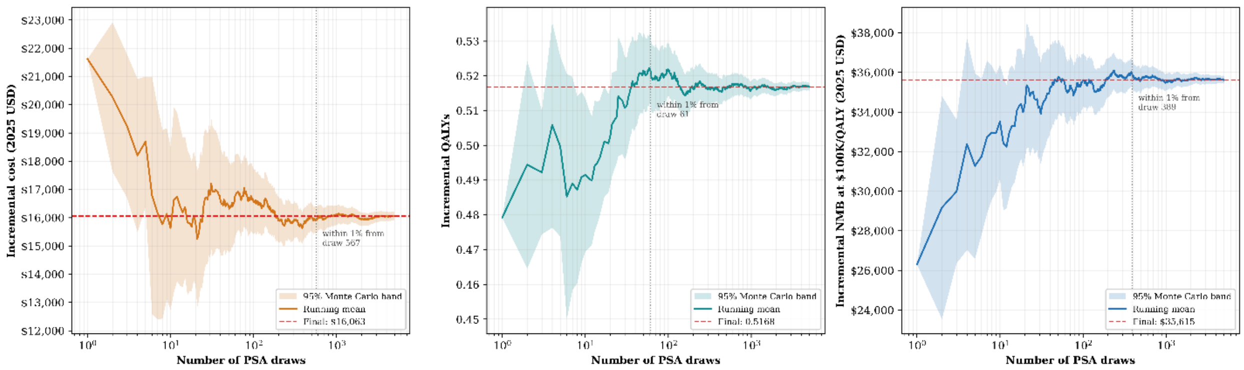

For both total costs and QALYs, the relative root-mean-square difference from the estimate based on all 50 replications declined steadily as additional replications were included and closely followed the expected square-root reduction in simulation error (**Figure E2**).

**Figure E2. Internal convergence across stochastic replications.** The reference curve shows the expected decline proportional to $\sqrt{1/n - 1/50}$, where $n$ is the number of accumulated replications. QALYs = quality-adjusted life years, PSA = probabilistic sensitivity analysis, reps = replications.

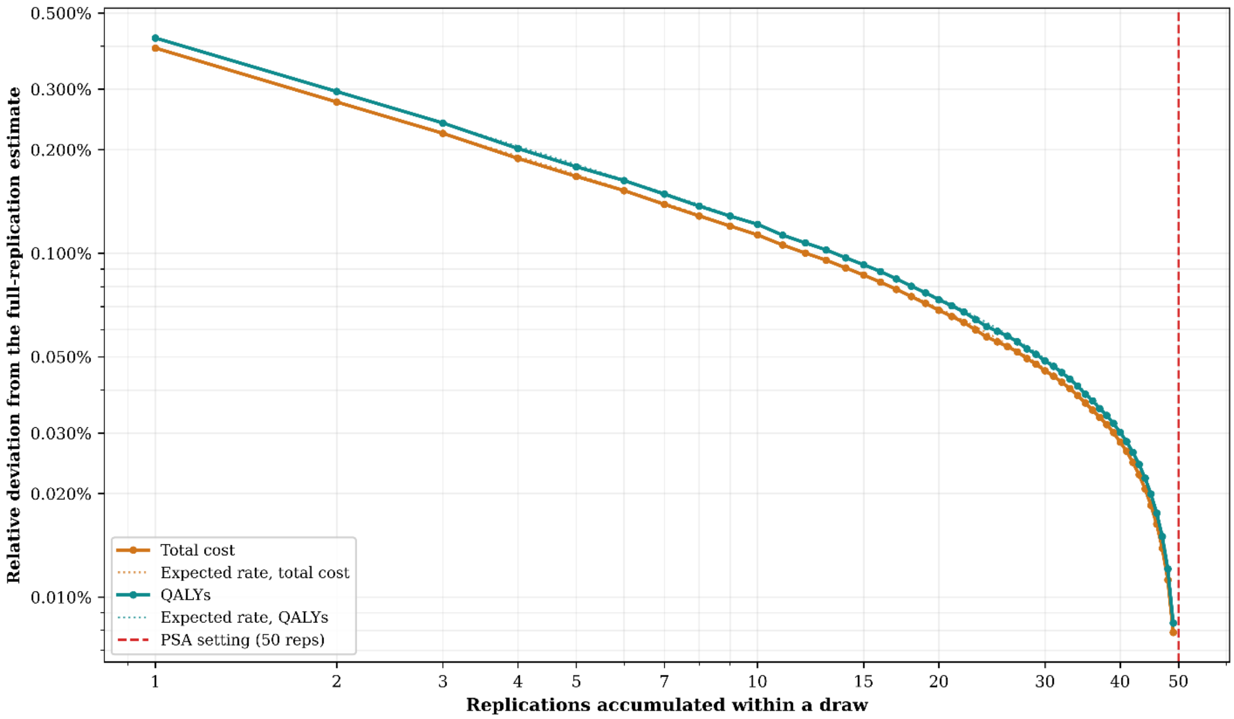

Relative deviation from the full-replication estimate
0.500%
0.300%
0.200%
0.100%
0.050%
0.030%
0.020%
0.010%
Replications accumulated within a draw
1
2
3
4
5
7
10
15
20
30
40
50
Total cost
Expected rate, total cost
QALYs
Expected rate, QALYs
PSA setting (50 reps)

### Appendix F. Cost-Effectiveness Across Intervention Scenarios

In the base case, the housing intervention generated a mean gain of 0.517 QALYs at an incremental cost of $16,063 per person. Doubling permanent housing costs increased incremental cost to $41,612 while leaving QALY gains unchanged. Higher utility while unhoused reduced the incremental QALY gain to 0.472 while leaving incremental costs unchanged (**Figure F1**).

**Figure F1. Probabilistic Cost-Effectiveness Planes.** Each point represents one paired probabilistic sensitivity analysis draw comparing the housing intervention with the status quo. Diamonds indicate mean incremental costs and quality-adjusted life years (QALYs).

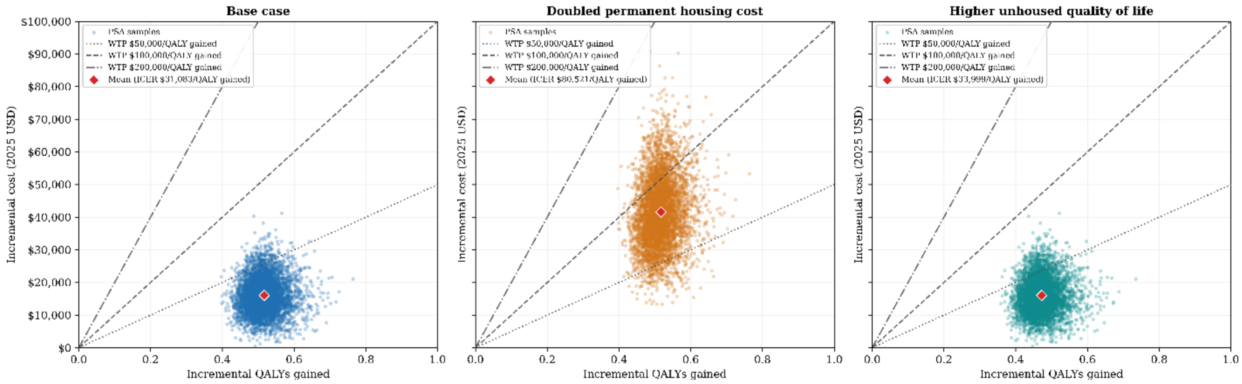

The willingness-to-pay values (at which expected incremental NMB crossed zero) were $31,083 per QALY gained in the base case, $80,521 with doubled permanent housing costs, and $33,999 with higher unhoused utility. At a willingness-to-pay threshold of $100K per QALY gained, the mean incremental NMB was $35,615, $10,066, and $31,182, respectively (**Figure F2**).

**Figure F2. Incremental Net Monetary Benefit (NMB) Across Willingness-to-Pay Thresholds.**

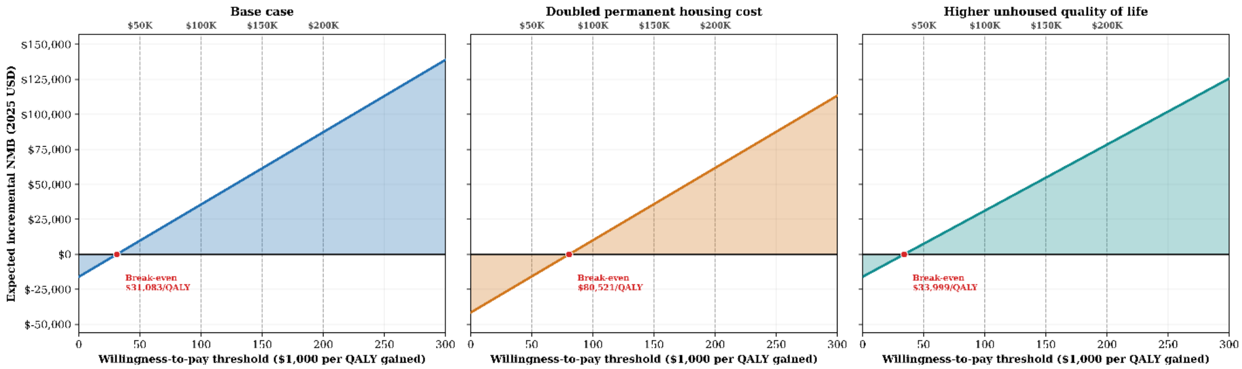

Mean incremental NMB increased with greater intervention achievement of the housing intervention under both cost scenarios (**Figure F3**). In the base case, the mean incremental NMB increased from $8,414 (95% UI, $5,307–$11,970) at 20% achievement of the housing intervention, to $16,098 ($10,212–$22,373) at 40%, $23,133 ($14,767–$32,143) at 60%, $29,620 ($18,952–$40,995) at 80%, and $35,615 ($22,773–$49,148) at 100%. With doubled permanent housing costs, the corresponding incremental NMB estimates were $2,339 (95% UI, -$3,044–$7,181), $4,499 (-$5,714–$13,556), $6,488 (-$8,231–$19,505), $8,343 (-$10,438–$25,003), and $10,066 (-$12,555–$30,022), respectively.

**Figure F3. Housing Intervention Effectiveness. Incremental Net Monetary Benefit (NMB) Per Person at a Willingness-to-Pay Threshold of $100,000 Per QALY Gained.**

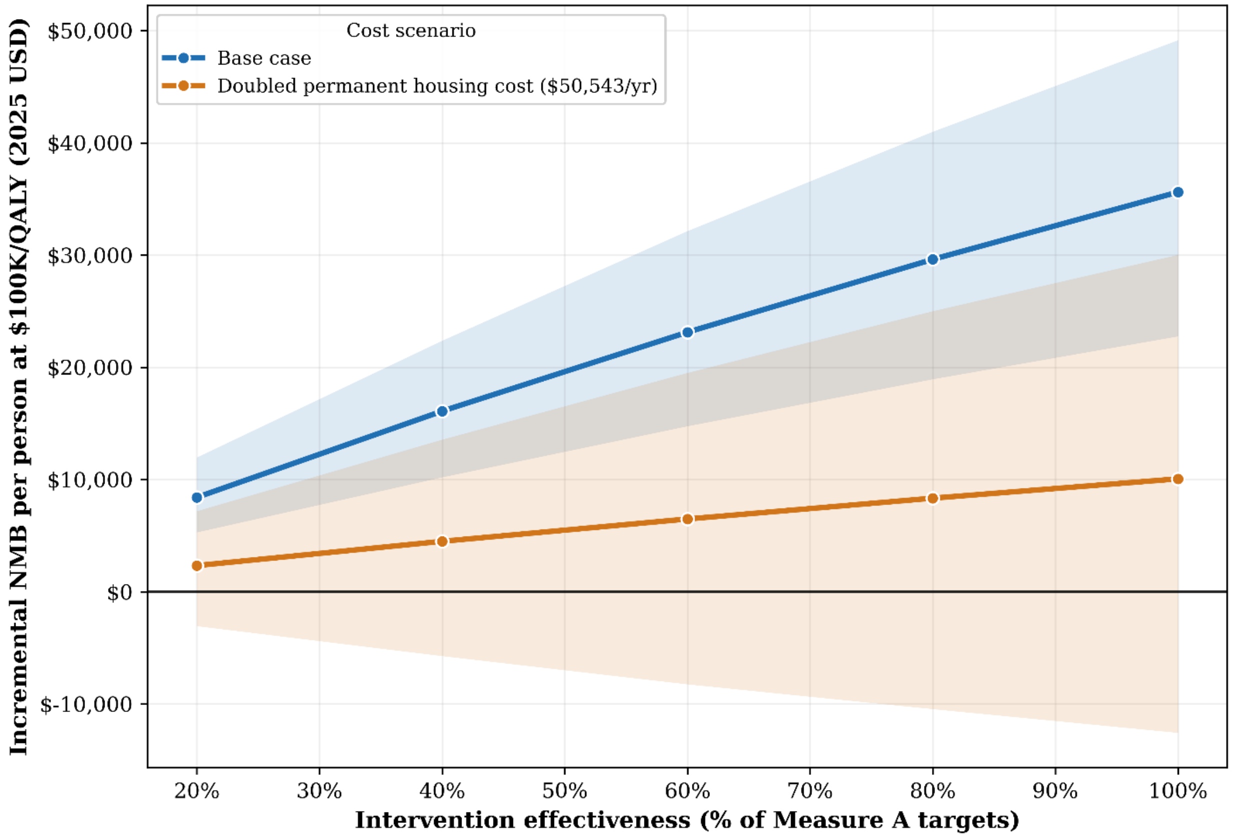
